# Mapping neurodevelopmental diversity in executive function

**DOI:** 10.1101/2023.06.15.23291392

**Authors:** Silvana Mareva, The CALM Team, Joni Holmes

## Abstract

Executive functions can be conceptualised as either a set of higher-order cognitive skills that enable us to engage in flexible thinking and regulate our thoughts and behaviours, or as the ability to integrate knowledge, beliefs, and values when applying cognitive control in everyday situations. These two perspectives map onto the ways in which executive function is measured in childhood – using either structured laboratory tasks or ratings of everyday behaviours. Differences in executive functioning are associated with neurodevelopmental differences, but evidence for associations between specific profiles of executive function and specific neurodevelopmental conditions is mixed. In this study, we adopt a data-driven approach to identify common profiles of executive function in a transdiagnostic sample of 566 neurodivergent children, using both performance and rating-based measures of executive function. Three profiles of executive function were identified: one had consistent difficulties across both types of assessments, while the other two had inconsistent profiles of predominantly rating- or predominantly task-based difficulties. Children with these different profiles had differences in academic achievement and mental health outcomes and could further be differentiated from a comparison group of neurotypical children on both shared and profile-unique patterns of neural white matter organisation. Importantly, children’s executive function profiles were not directly related to diagnostic categories or to dimensions of neurodiversity associated with specific diagnoses (e.g., hyperactivity, inattention, social communication). These findings support the idea that there are separate domains of executive function and that the two types of assessment tapping these functions are dissociable and provide non-redundant information related to neurodevelopmental differences. These findings advance our understanding of executive function profiles in neurodivergent populations and their relationship to behavioural outcomes and neural variation.

## Introduction

The transdiagnostic revolution in neurodevelopmental conditions emphasises a move away from diagnostic systems towards identifying broad dimensions that characterise neurodiversity (Astle et al., 2021). One such dimension, which captures characteristics associated with multiple neurodevelopmental outcomes (e.g., Bloemen et al., 2018) and can be a barrier to learning (Holmes et al., 2020) is executive function. The term executive function is used widely to describe the processes supporting the volitional control of cognition and behaviour. In this study, we adopt a transdiagnostic data-driven approach to delineate profiles of executive function among a large neurodevelopmentally neurodivergent sample using measures designed to capture two conceptually distinct aspects of executive function: performance-based tasks designed to tap higher-order cognitive skills (Miyake et al., 2000), and behaviour ratings suited to capturing the application of cognitive control in everyday contexts (Doebel, 2020). We then explore whether and how these data-driven groups are associated with differences in neurodevelopmental diagnostic categories and dimensions, academic and mental health outcomes, and neural white matter organisation.

Executive function can be conceptualised in two ways that map broadly on to the ways in which they are measured (Malanchini et al., 2018; Toplak et al., 2013). The first views executive function as a set of higher-order cognitive skills that enable flexible thinking (Miyake et al., 2000). These abilities include shifting/switching, inhibition, working memory, planning, and sustained attention (Friedman & Miyake, 2017; Miyake et al., 2000; St Clair-Thompson & Gathercole, 2006a). They are typically indexed by accuracy or response times on performance-based tasks. These include memory span tasks that index working memory performance (e.g., Automated Working Memory Assessment, Alloway, 2007), tasks that require goal-oriented planning (e.g., Tower of London, Shallice, 1982), semantic incongruence or cognitive flexibility tasks that assess shifting/switching (e.g., Number-Letter sequencing, Delis et al., 2001; Stroop, Stroop, 1935), and tasks that require focused attention in the face of distraction over a lengthy periods that measure sustained attention and response inhibition (e.g., Continuous Performance Test, Conners, 1992). There are many other task-based measures of executive function, and multiple variants of the examples provided here (see Strauss et al., 2006 for a comprehensive list of task-based measures of executive function), but what they have in common is that they are administered individually under controlled experimental conditions requiring effortful cognitive control. In this way, therefore they can be viewed to reflect optimal rather than typical performance.

The second view of executive function integrates knowledge, beliefs, and values alongside cognitive, motor, and perceptual control, and makes explicit that these factors come together in the service of goals in specific settings (Doebel, 2020). It refers to the individual use of executive skills in particular situations, rather a general “capacity process” such as inhibitory control that is applied across all situations. To illustrate the distinction, Doebel (2020) uses the example of a child inhibiting the impulse to hit another child who has taken their toy. In this situation, the child brings to bear knowledge and beliefs about hitting another child (e.g., socially acceptable alternatives to hitting, beliefs about being scolded for engaging in hitting behaviour) when exerting inhibitory control. In this way, executive functions are viewed as inseparable from the task goals, context, and mental content that comes into play in specific situations. Understanding executive function as a concept that is tied to the activities a person is completing in their everyday activities mirrors the second way in which executive function is measured – using rating scales to index the ability to coordinate multiple processes in everyday problem-solving situations that draw on knowledge, beliefs, and values. Examples of these rating scales include the Behaviour Rating Index of Executive Function (BRIEF; Gioia et al., 2000) and the Working Memory Rating Scale (Alloway et al., 2009). These, and similar questionnaires, require observers to rate the frequency of everyday executive function behaviours in different contexts. They were originally developed to provide an ecologically valid measure of executive function that captures real-world functioning. Questionnaire responses typically reflect behavioural observations over extended periods of time, and may therefore reflect more trait-like or stable processes and provide a more accurate assessment of typical performance (Malanchini et al., 2018).

Two lines of evidence suggest that executive function in these two domains – cognitive processes or skills involved in goal-oriented behaviour in everyday situations – is dissociable. First, there is little convergent validity between the measurement types tapping each domain (Dang et al., 2020). If performance-based and rating-based measures of executive function were assessing the same underlying construct, they should be highly correlated. Yet concurrent and predictive validity studies consistently indicate that they are, at best, weakly correlated (e.g., Gerst et al., 2017; Nin et al., 2022; Soto et al., 2020; Tamm & Peugh, 2019; Toplak et al., 2013), and that these relationships remain small when using latent variable approaches (Snyder et al., 2021) and/or when controlling for mono-method bias (e.g., that performance-based measures predict other performance-based measures, and rating-based measures predict other rating-based measures Soto et al., 2020). In a review, Toplak and colleagues (2013) concluded that the two types of measurement capture different information, with rating-based measures capturing goal-oriented successes in typical everyday settings and performance-based measures capturing processing-efficiency under optimal conditions.

Second, they make independent contributions to clinical, academic, and life outcomes. Difficulties with executive function are associated with multiple neurodevelopmental conditions, including attention deficit hyperactivity disorder (ADHD), specific learning difficulties, and autism (e.g., Benallie et al., 2021; Holmes et al., 2014; Loe & Feldman, 2007; McClain et al., 2022; Willcutt et al., 2005), they can be a barrier to learning (Holmes et al., 2020; Peng & Fuchs, 2016; Soto et al., 2021; Yeniad et al., 2013), and are also linked to poor mental health outcomes (e.g., Bloemen et al., 2018). Substantial evidence suggests that the different measurement methods corresponding to each domain of executive function make unique and separable contributions to these outcomes (Gerst et al., 2017; Soto et al., 2020). In terms of academic achievement, both rating- and performance-based measures predict outcomes, but the relationships are typically stronger and more consistent for performance-based measures (Gerst et al., 2017; Malanchini et al., 2018; Soto et al., 2020). Similarly, for mental health outcomes, rating- and task-based measures of executive function explain independent variance in both internalising and externalising difficulties (Eisenberg et al., 2019; Ellingson et al., 2019; Friedman et al., 2020; Friedman & Gustavson, 2022), with the stronger relationships typically reported between rating-based measures and mental health (Friedman et al., 2020).

The correspondence between ratings of executive function behaviours and performance on cognitive tests is also low in neurodivergent groups. For example, in individuals with ADHD, the executive function profiles captured on performance-based and rating-based measures have little overlap (Biederman et al., 2008). Moreover, both types of assessments have their own distinct contribution to predicting occupational outcomes in this population (Barkley & Murphy, 2010, 2011). In a transdiagnostic study, Williams and colleagues (2022) found that a substantial proportion of a large sample of neurodivergent children had what they termed as an inconsistent executive function, defined as difficulties reported on rating-based assessments of executive function that were not evident on performance-based tasks.

### Current study

The aim of the current study was to adopt a transdiagnostic approach to delineate profiles of executive function among a large neurodevelopmentally neurodivergent sample. Previous studies exploring executive function in neurodivergent children have typically either contrasted executive function performance between groups (e.g., those with a diagnosis compared those with a different, or no, diagnosis) across a range of performance-based measures (e.g., Geurts et al., 2004; Holmes et al., 2014), explored how well executive function rating scales and performance tests predict outcomes separately (e.g., Gerst et al., 2017; Soto et al., 2020), or tested relationships between different types of executive function measurement (e.g., Nin et al., 2022; Tamm & Peugh, 2019). Here, we adopted a different approach to explore individual profiles of executive function across both performance- and rating-based measures that were designed to tap into the cognitive process-based and situational goal-oriented conceptualisations of executive function. We used a self-organising map algorithm to map the multidimensional space of a broad range of performance and rating-based assessments of executive function, representing how children group together based on their executive function profiles. We then used data-driven clustering to delineate subgroups of children presenting with different executive function profiles and explored how these related to the two dissociable conceptualisations of executive function. We also investigated whether the groups differed in terms of neurodevelopmental diagnostic categories (ADHD, autism, etc.) and dimensions that capture neurodiversity (hyperactivity, inattention, social communication). Finally, we explored how these groups related to learning, mental health, and neural white matter organisation. We focussed on white matter organisation because there is a substantial literature supporting its links to both executive function (Bathelt et al., 2018; Baum et al., 2017) and neurodevelopmental conditions and dimensions (Ameis et al., 2016; Beare et al., 2017). We applied this approach to data from a transdiagnostic sample that included a range of children who had been identified as having additional needs by health and education practitioners, irrespective of their diagnostic status, consistent with the notion that diagnostic-based studies fail to capture broad populations of neurodivergent children (Astle et al., 2021). The study was fully exploratory, and we did not formulate a hypothesis about the number of groups or the phenotypic and neural features that would differentiate them. Instead, we used a data-driven approach to address the following questions: 1) can we identify subgroups of children with different profiles of executive function; 2) do groups differ in terms the two dissociable conceptualisations of executive function; and 3) how do these groups relate to neurodevelopmental diagnostic characteristics, academic and mental health functioning, and neural white matter organisation?

## Method

### Participants

Participants were drawn from the Centre for Attention, Learning, and Memory (CALM) cohort. Recruitment details and testing procedures are described in the study protocol (Holmes et al., 2019). Broadly, children aged 5-18 years were referred to the study by health and educational professionals for difficulties with attention, learning, and/or memory regardless of diagnostic status. Participants aged 8 years and above were included in the current study. Younger children were excluded because some of the executive function tasks were not standardised for children under 8 years and were therefore not administered to them. The sample for this study included 566 children (45% diagnosed) with an average age of M = 10.55 years, SD = 2.02 (N_boys_ = 380, N_girls_ = 186). The most common diagnoses were ADHD (N=156), Autism (N=46), and dyslexia (N= 47). Ethical approval was obtained by the National Health Service (REC: 13/EE/0157). Parents/caregivers gave written consent and children gave verbal assent to participate.

### Measures

Children completed a range of assessments of learning and cognition and a questionnaire asking about their mental health. Parents/caregivers completed questionnaires about the child’s behaviour, communication skills, and mental health. All assessments were administered by a trained researcher at the CALM clinic, following standardised administration procedures documented in the test manuals. Task details and administration procedures are available in the study protocol and summarised briefly below.

### Executive Function

#### Task-based measures

Tasks tapping executive functioning were chosen from the broader neuropsychological battery used in the CALM study (Holmes et al., 2019). These included four tasks from the Automated Working Memory Assessment (AWMA; Alloway, 2007), tapping verbal and visuospatial short-term/working memory: forward digit recall, backward digit recall, dot matrix, and Mr X. Two tasks from the Delis-Kaplan Executive Function System test battery (D-KEFS, Delis et al., 2001) were included: the Tower task, commonly used to assess planning skills, and the Trail-making number-letter sequencing task, a Stroop-like task indexing shifting. Two tasks from the Test of Everyday Attention for Children 2 (TEA-Ch2, Manly et al., 2016) were also included: the switching task, Reds, Blues, Bags and Shoes (RBBS), and the Vigil task tapping sustained attention.

#### Rating-based measures

Parents/caregivers completed the BRIEF (Gioia et al., 2000), an 80-item rating scale covering eight domains: Inhibition, Shifting, Emotional control, Initiation, Working memory, Planning, Organisation, and Monitoring.

### Learning

The Word Reading and Numerical Operations subtests of the Wechsler Individual Achievement Test II (WIAT-II; Wechsler, 2005) were used to assess children’s reading and mathematical skills.

### Mental health

The total anxiety and depression score from the Revised Child Anxiety and Depression Scale (RCADS, Chorpita et al., 2005) was used to index child-reported internalising difficulties. This questionnaire was introduced into the CALM protocol mid-way through the study, meaning data were only available for 269 participants.

### Diagnostic categories and dimensions of neurodiversity

Neurodevelopmental diagnoses were reported by the referring professional and confirmed by parents/ caregivers at the time of referral to the CALM study. Ratings on dimensions of neurodiversity that are core features of neurodevelopmental conditions that were included in the CALM protocol were included in the current study. These were ratings provided by the child’s parent / caregiver using the following scales: inattention and hyperactivity/impulsivity (dimensions of ADHD measured by the Conners-3 Parent Short Form Rating Scale (Conners, 2008); communication skills (dimension of developmental language disorder measured by the Child Communication Checklist-2, CCC-2, Bishop, 2003), and social communication and interests (core dimensions of autism, measured by the CCC-2, Bishop, 2003).

### MRI data acquisition and pre-processing

All participants were invited to participate in an optional magnetic resonance imaging (MRI) session. T1-weighted volume scans were acquired using a whole-brain coverage 3D Magnetization Prepared Rapid Acquisition Gradient Echo (MP RAGE) sequence acquired using 1 mm isometric image resolution. Diffusion scans were obtained using echo-planar diffusion-weighted images with an isotropic set of 68 noncollinear directions. Whole brain coverage was based on 60 contiguous axial slices and isometric image resolution of 2 mm. Echo time was 90 ms and repetition time was 8,500 ms. QSIPrep 0.13.0RC1 (based on Nipype 1.6.0 Gorgolewski et al., 2011) was used for MRI pre-processing and reconstruction. Whole-brain white matter connectivity matrices (i.e. connectomes) were constructed for each child based on the Brainnetome atlas (Fan et al., 2016). For each pairwise combination of regions (N = 246), the number of streamlines intersecting them was estimated and transformed to a 246 x 246 streamline matrix.

Only a subsample of the children referred to CALM agreed to participate in the neuroimaging session (N = 248, Age scan = 10.92, SD = 2.07, 70% male). The neuroimaging cohort also included 77 children who formed a comparison group. These children (N = 77, Age scan =10.75, SD = 2.01) were not referred by health and educational professionals but were recruited from the same schools attended by those who were referred. This non-referred / neurotypical group was included in the neuroimaging analyses, because unlike the cognitive and behavioural assessments, neuroimaging metrics do not allow for straightforward interpretation of age-typical levels of functioning.

### Analysis Plan

Analyses were conducted in four steps: 1) missing data estimation; 2) self-organising map (SOM) and data-driven clustering; 3) comparison of profiles across executive function, neurodevelopmental, academic, and mental health functioning based on a series of chi-squares and t-tests; 4) pairwise comparisons and partial least squares discriminant analysis (PLSDA) to explore patterns of neural white matter organisation that distinguish the data-driven groups from the neurotypical participants who were included in the neuroimaging cohort. Details for each step of the analysis are outlined below.

### Missing data

Missing data were estimated only for the assessments used in the SOMs. For all other assessments used in the validation analyses, missing data was not estimated to ensure it was independent and suitable for external validation. Missingness across executive function assessments used in the SOMs ranged from 0.7% to 25.3%. Data was complete for 59% of participants. Boys and girls were equally likely to have missing data (χ2 = 0.22, p = 0.64) and so were diagnosed and undiagnosed participants (χ2 = 0.03, p = 0.87). Missing data was estimated via the random forest nonparametric imputation procedure implemented in R package missForest (Stekhoven & Bühlmann, 2012). Participant sex and age were also added to the imputation model to improve estimates.

### Self-organising maps (SOMs)

Self-organizing maps (SOMs) are a type of artificial neural network suitable for analysing high-dimensional data in a lower-dimensional space. SOMs use unsupervised learning to group similar input data together and create a topological map, where neighbouring nodes correspond to similar input patterns. Here we provide a brief conceptual overview of the implementation used in the current analysis, for more information about the SOM algorithm, see Wehrens & Buydens, (2007). SOMs consist of a predefined number of nodes laid out on a grid – in this case, hexagonal nodes on 10 by 10 grid, where each node in the grid corresponded to a weight vector with the same dimensionality as the input data (N =16, corresponding to the number of executive function measures used to train the SOMs). The training of the map began with a subset of the data randomly assigned to the units. The process was repeated in 1000 iterations, and each time the weights of a best-matching unit (i.e., the node most alike the current training object based on the least shared Euclidean distance), and its neighbouring nodes, were updated to become more similar to the input data. In this way, neighbouring nodes became more alike. At the end of the training process, the weight vector for each node reflected the executive function scores of the children for whom that node was the best matching unit, with neighbouring nodes having similar weights. The resulting map represented a model of the multidimensional executive function data on which the SOM was trained, whereby children with similar profiles across the 16 executive function assessments “sat” closer in space. Age-standardised scores were used in these analyses. These were a combination of T-, scaled and standard scores, so they were converted to age-referenced z-scores to be on the same scale (score of 0 corresponded to age-typical performance, 1 = 1 SD above the age-expected mean, and −1 represented 1 SD below the age-expected mean). SOMs were trained using the *R* package kohonen (Wehrens & Buydens, 2007).

### Data-driven clustering

The SOM maps the data in a continuous two-dimensional plane of nodes, where space indicates similarity. Therefore, carving the map into sections should yield groups with relatively homogenous profiles who are different from children sitting elsewhere on the map. Given the unsupervised nature of the SOMs, there is no pre-defined rationale for how many sections the map should be divided into. To identify the optimal number of groups, we used a data-driven method in which the nodal weight values from the SOM were submitted to a data-driven clustering. Following good practice recommendations, before clustering, the nodal weights were reduced using uniform manifold approximation and projection (Bathelt et al., 2021; Dalmaijer et al., 2022). The optimal number of clusters was chosen based on a consensus approach implemented in the R-package NbClust (Charrad et al., 2014). This method uses 30 indices for determining the number of clusters and converges on the best clustering scheme across the different results obtained by combinations of the number of clusters, distance measures, and clustering methods. Once the nodes were grouped according to the similarity of their weights, we identified children assigned to each group of nodes. This provided us with clusters of children based on nodes they were assigned to in the original mapping. To check the clustering, each cluster distribution was plotted on the original map with the expectation that all cluster members ought to sit on neighbouring nodes within the original map.

### Cluster characterisation & comparisons

To characterise the data-driven groups we compared their average scores on each of the assessments used in the SOM in a series of t-tests. False discovery correction was used to handle multiple comparisons. We also validated the data-driven clusters by testing whether group differences generalised to data not included in the SOM – in this case, learning assessments and self-reported mental health difficulties, neurodevelopmental diagnostic categories, and parent-reported dimensions of neurodiversity. For continuous data, t-tests were conducted using false discovery rate as a correction for multiple comparisons, and for categorical data, chi-square tests were used (e.g., to compare the number of children with a particular diagnosis in each subgroup relative to the overall distribution across the whole sample).

### Neuroimaging comparisons

The neuroimaging analyses explored three levels of white matter connectome organisation: global, module, and regional hubs. For the first two, we used series of non-parametric comparisons contrasting the non-referred neurotypical group to the data-driven groups using false-discovery rate correction. This approach was favoured as it allowed us to explore both shared and subgroup-unique differences relative to the comparison group with a minimum number of comparisons. To explore differences in regional hub organisation we used a PLSDA (Brereton & Lloyd, 2014), aiming to derive components of regional hubs that best explain group membership across the data-driven profiles and the non-referred neurotypical sample. Prior to analyses, the effects of age and in-scanner motion (average frame displacement) were regressed from each metric using a robust estimation approach.

At the global level, we focused on connectome efficiency and the average participation coefficient. Global efficiency describes the potential for information exchange in the connectome and is the average inverse distance from any one brain region to another (Sporns et al., 2007). The participation coefficient measures the proportion of possible connections each region has with regions from other modules. It is considered an index of modular segregation, with lower values indicating more segregation: regions with a high participation coefficient have strong connections to many modules, while regions with a low participation coefficient have strong connections to fewer modules (Baum et al., 2017). A global participation coefficient was derived by averaging across the 246 regions. These metrics were chosen because they have been previously linked to improved performance on tests of executive function in developing populations (Baum et al., 2017; Koenis et al., 2015).

To derive module organisation, we grouped the 246 regions of the Brainnetome atlas a priori into their corresponding functional networks as defined by Yeo et al. (2011). Subcortical nodes were also grouped together. This modular parcellation is based on independent functional data but has been previously shown to be a good representation of the organisation seen in the white matter connectome in developing populations (Baum et al., 2017). We then estimated modular strength for each one of the 7 networks and the subcortex. Significantly different results were followed-up to explore whether differences were driven by changes in within-module connectivity, between-module connectivity, or both.

Finally, PLSDA was chosen as a tool to explore links between regional connector hub strength and group membership. PLSDA is a statistical technique used to find relationships between two sets of variables: a predictor set and a response set. In our case these were connector hub strength and group membership respectively. The goal of PLSDA was therefore to identify the linear combination of connector hubs that best explains the variation in group membership. We focused on connector hubs because they play an important role in network specialisation across development and support the development of executive function (Baum et al., 2017; Jones et al., 2021; Zink et al., 2021). In accordance with prior work, connector hubs were defined as regions which were above the 70th percentile on both betweenness centrality and the participation coefficient (Jones et al., 2021). The analysis was implemented in the R package mixOmics (Rohart et al., 2017). The model was evaluated by using 5-fold cross-validation repeated 50 times and the number of components to retain was chosen based the balanced error rate metric, accounting for the uneven number of participants across groups (N cluster 1 = 95, N*c*luster2 =74, N cluster3 =79). The contribution of each hub (i.e., component loadings) to the PLSDA components was then evaluated using a bootstrap procedure (N= 1000), where a Procrustes rotation was applied to align the factors across iterations (Krishnan et al., 2011). This was done to test whether the bootstrapped confidence interval passed zero, and thus to establish which hubs reliably load on the PLSDA components. Group differences in component scores were compared using a non-parametric permutation procedure testing whether they significantly differed from chance when group labels were permuted 1000 times.

## Results

### SOMs and Clustering

Performance across all measures used in the SOM are presented in Table 1, and correlations between the measures in Figure 1. The outcomes of the trained SOM are presented in Figures 2, showing how the SOM represents the values for each weight vector (i.e., the weights that correspond to each individual executive function measure) across the grid of nodes. Each panel shows the distribution of weights for a different executive function measure. Data-driven consensus clustering applied to the SOM suggested that a three-cluster solution was optimal (the map separated by cluster can be seen in Figure 3).

**Figure 1.**
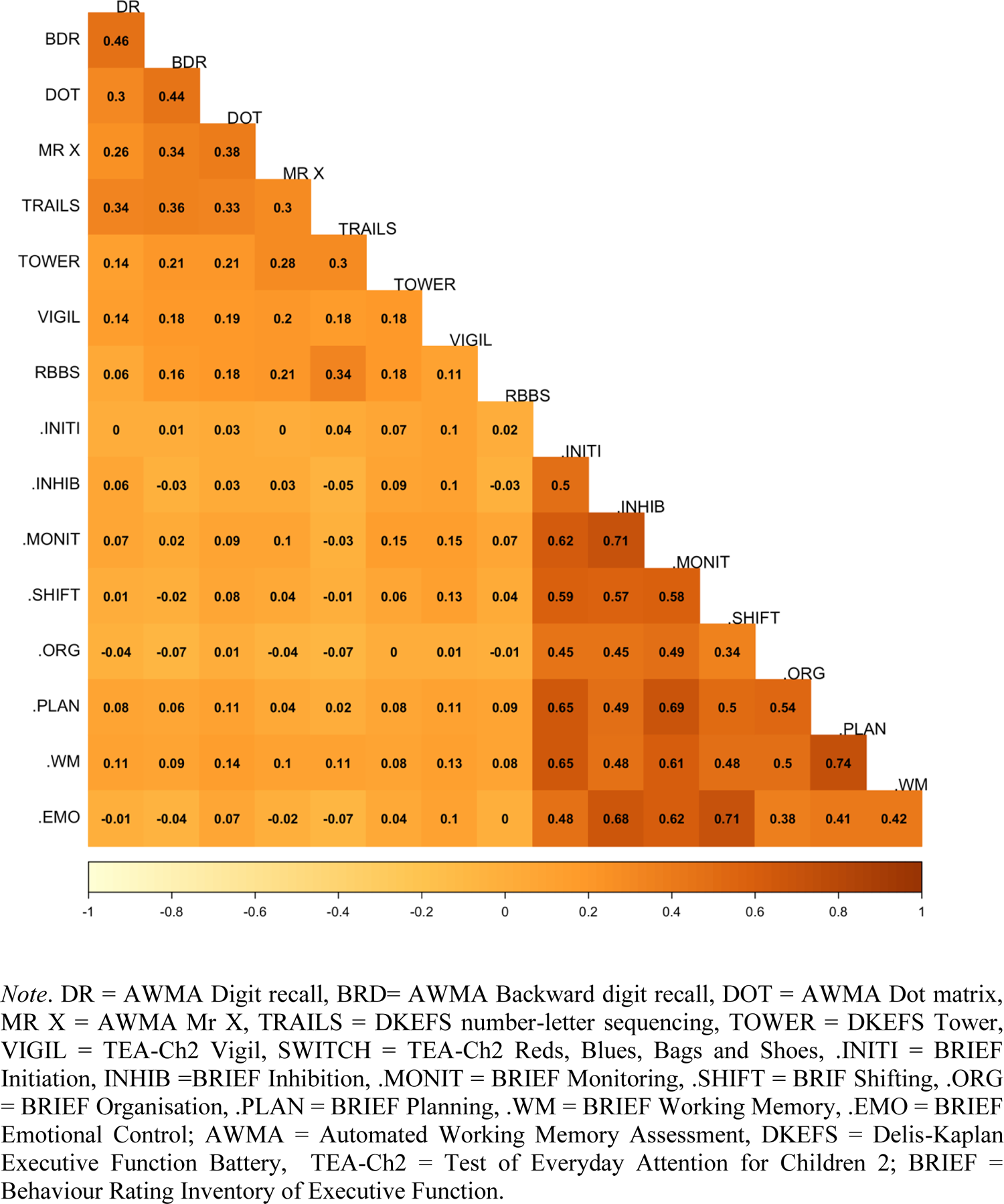
Pearson correlations across all norm-referenced age-corrected executive function assessment scores used in the self-organising map

**Figure 2.**
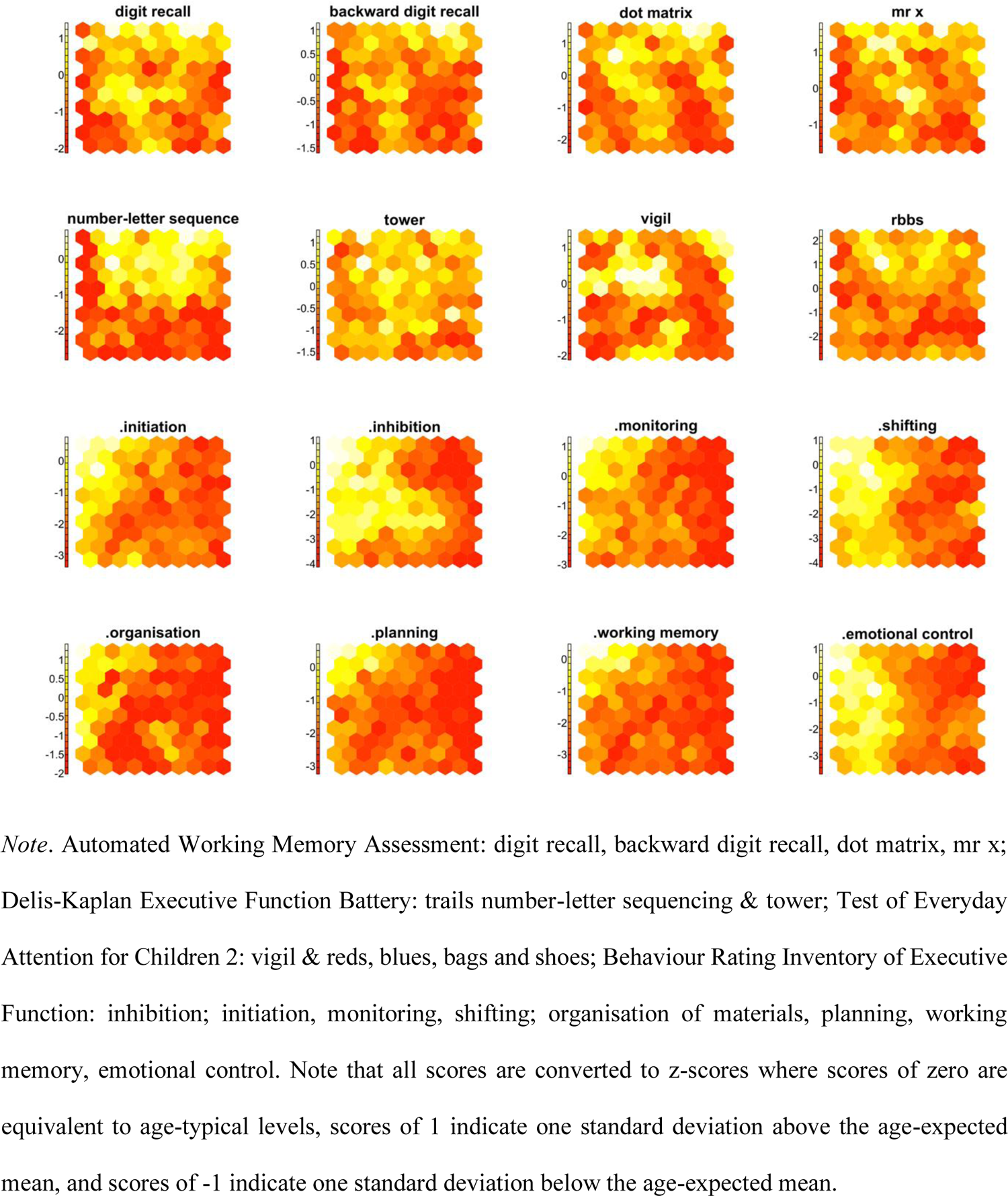
Weight distributions from the self-organizing map, split by assessment. For each assessment, the map depicts high weights (i.e., relatively stronger performance/ratings) as yellow hexagons and low weights (i.e., below age-expected performance/rating) as red hexagons.

**Figure 3.**
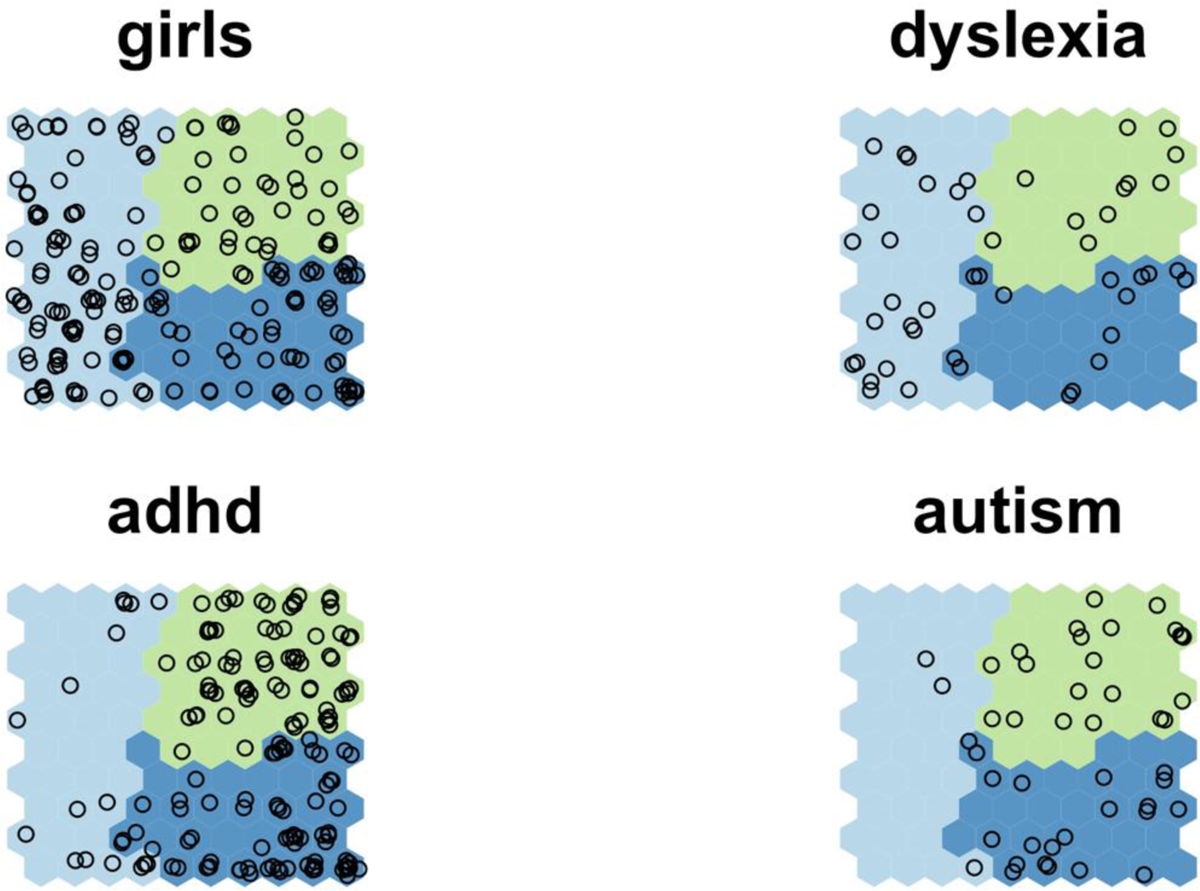
Distribution of sexes and different diagnoses across data-driven groups.

**Table 1.** Norm-referenced age-corrected means and standard deviations for the whole sample across all measures used in the self-organising map and clustering.

|  | Mean | SD |
| --- | --- | --- |
| AWMA: Digit recall | 91.62 | 15.59 |
| AWMA: Backward digit recall | 91.39 | 11.46 |
| AWMA: Dot matrix | 90.02 | 15.60 |
| AWMA: Mr X | 95.90 | 14.43 |
| DKEFS: Trails: number-letter sequencing | 6.14 | 3.44 |
| DKEFS: Tower | 9.41 | 2.28 |
| TEA-Ch2: Vigil | 8.26 | 3.20 |
| TEA-Ch2: Reds, Blues, Bags and Shoes | 7.69 | 3.36 |
| BRIEF: Initiation | 66.46 | 10.43 |
| BRIEF: Inhibition | 66.16 | 15.15 |
| BRIEF: Monitoring | 65.51 | 10.71 |
| BRIEF: Shifting | 68.24 | 14.52 |
| BRIEF: Organisation | 60.08 | 9.82 |
| BRIEF: Planning | 70.41 | 9.57 |
| BRIEF: Working memory | 73.28 | 9.60 |
| BRIEF: Emotional control | 64.60 | 13.30 |

*Note.* AWMA = Automated Working Memory Assessment; DKEFS = Delis-Kaplan Executive Function Battery; TEA-Ch2 = Test of Everyday Attention for Children 2; BRIEF = Behaviour Rating Inventory of Executive Function.

### Cluster Characterisation

The group means for each of the three data-driven groups on the executive function assessments used in the SOM are shown in Figure 4. One group (Cluster 1, N = 204) was characterised by close to age-expected levels across the rating-based assessments, with fewer difficulties compared to the other two clusters. Their performance on the task-based measures was poorer than their ratings, and below age-expected levels. Overall, they had better task-based performance compared to children in Cluster 2 and poorer performance than those in Cluster 3. Based on the discrepancy between their rating- and task-based performance, children in Cluster 1 are referred to as having an inconsistent profile of predominantly task-based difficulties.

**Figure 4.**
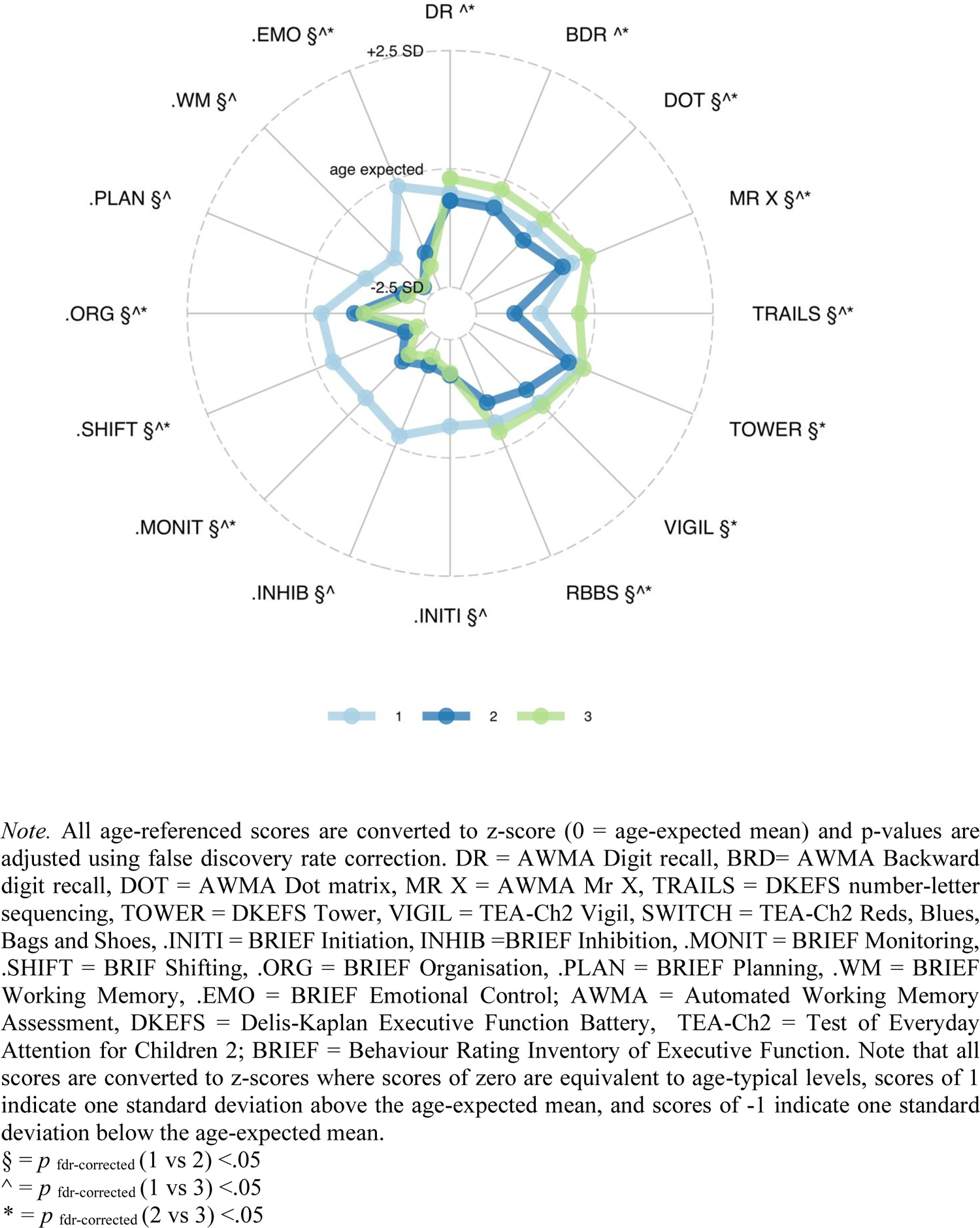
Executive function scores across all assessments used in the self-organising map for each one of the data-driven groups.

A second group, Cluster 2 (N=187), was characterised by widespread difficulties across both the rating- and task-based executive function assessments and was therefore labelled as having consistent difficulties. The third group, Cluster 3 (N = 175), included children with the opposite profile to those in Cluster 1. They had an inconsistent profile with close to age-expected levels of performance on the task-based measures with pronounced difficulties on the rating-based assessments that were indistinguishable from those of the children in Cluster 2 who had a profile of consistent difficulties. Children in Cluster 3 are referred to as having an inconsistent profile of predominantly rating-based difficulties.

There were significant differences in the average age of children in each of the three Clusters (F = 9.56, p <.01): children in Cluster 1 who had an inconsistent profile of predominantly task-based difficulties were on average younger than children in Cluster 2 who had a consistent profile of difficulties (t= −3.50, p <.001, mean difference = 0.71 years) and children in Cluster 3 who had predominantly rating-based difficulties (t = −3.06, p <.002, mean difference = 0.62 years). There was no significant difference in age between children in Clusters 2 and 3. To explore whether child sex was related to group membership, while accounting for the overrepresentation of boys in the sample as a whole, a series of chi-square tests were conducted to compare the sex distribution observed in each cluster to the sex distribution of the whole sample (Figure 4). Girls were overrepresented in Cluster 1 (profile with predominantly task-based difficulties, *χ^2^*= 5.66, *p*=.02), and underrepresented in Cluster 3 (profile with predominantly rating-based difficulties, *χ^2^*= 7.94, *p*=.005). The distribution of boys and girls in Cluster 2 (consistent difficulties group) matched that of the whole sample (*χ^2^*= 0.06, *p*=.81), suggesting that boys and girls were equally likely to have this profile.

### Cluster validation

To validate differences between the three clusters, their performance was compared across measures of learning, mental health, and dimensions of neurodiversity, and the distribution of children with diagnosed neurodevelopmental conditions in each cluster was compared to the distribution across the whole sample. Children in Cluster 3 (inconsistent profile of predominantly rating-based difficulties) had significantly better performance across measures of reading and maths relative to children in the other two clusters (Figure 5). Those in Cluster 2 (consistent difficulties profile) had significantly lower performance on the maths measure relative to children in Clusters 1 and 3, but their reading performance was indistinguishable from that of children in Cluster 1 who had predominantly task-based difficulties. For mental health, children in Cluster 3 (predominantly rating-based difficulties) had significantly elevated levels of anxiety and depression relative to children in the other two clusters; their scores were not significantly different from one another (see Figure 5).

**Figure 5.**
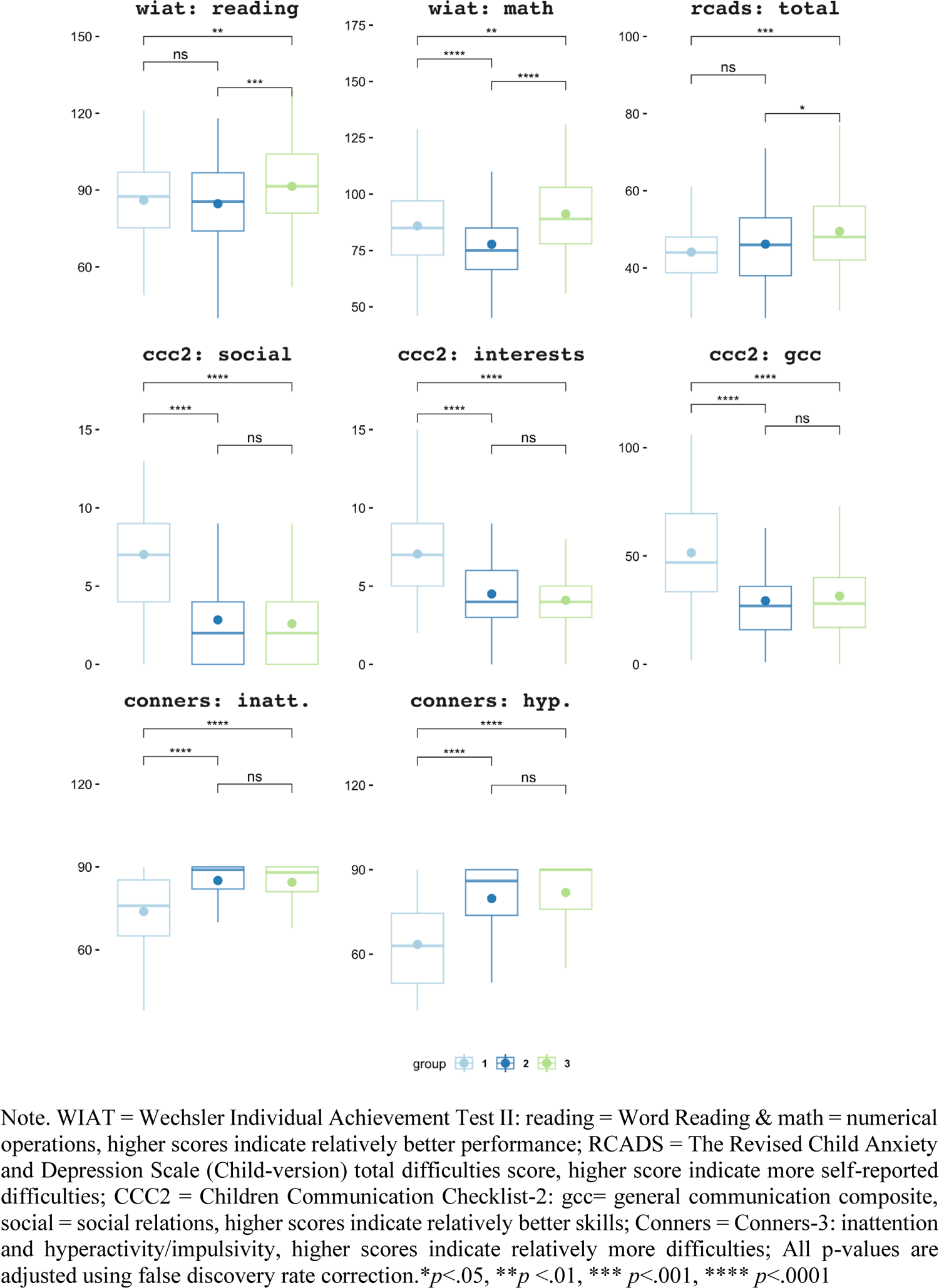
Results of pairwise comparisons of the data-driven groups across measures of learning, self-rated mental health, and patent-rated neurodevelopmental diagnostic dimensions.

Autistic participants and those with an ADHD diagnosis were underrepresented in Cluster 1, the group with predominately task-based difficulties (ADHD: *χ^2^*= 39.73, *p*<.001; Autism: *χ^2^*= 12.11, *p*<.0001), but were represented in similar proportions in the other two clusters. It should be noted that 41% of autistic participants also had an ADHD diagnosis, meaning separate conclusions cannot be drawn for the two conditions. Children with a dyslexia diagnosis were present in each cluster, and there was no relationship between dyslexia diagnosis and group membership (ps >.05). A similar pattern emerged across dimensions associated with neurodevelopmental conditions. Across the five dimensional measures (inattention, hyperactivity/impulsivity, general communication skills, social communication and interests), children in Cluster 1 (predominantly task-based difficulties) had significantly fewer difficulties than those in Clusters 2 and 3, who did not differ significantly from one another. In other words, there was no clear one-to-one correspondence between data-driven group membership and neurodevelopmental diagnostic dimensions. Instead, relative to the group with primarily task-based difficulties (Cluster 1), the groups with consistent and predominantly rating-based difficulties (Clusters 2 and 3) included more children with diagnosed neurodevelopmental conditions and experienced more neurodevelopmental difficulties on dimensional measures.

### Neuroimaging comparisons

At the global level (Figure 6), children with consistent difficulties across rating- and task-based measures (Cluster 2) and those with mainly rating-based difficulties (Cluster 3) had significantly lower global connectome efficiency compared to the non-referred neurotypical group. The three data-driven groups did not differ significantly from the non-referred comparison group in average connectome participation coefficient. Looking at modular organisation, Clusters 2 (consistent difficulties) and 3 (predominantly rating-based difficulties) had similar patterns of neural organisation that differentiated them from the non-referred comparison group: they both showed reduced modular strength in the limbic network and the subcortex (Figure S1). The group with primarily task-based difficulties (Cluster 1) showed the same pattern of reduced modular strength in the limbic network and the subcortex but was also characterised by increased strength in somatomotor and ventral attention modules relative to the comparison sample (Figure S1). Follow-up analysis suggested that all these differences were driven by differences in between - rather than within - modular connection strength (Figure 6).

**Figure 6.**
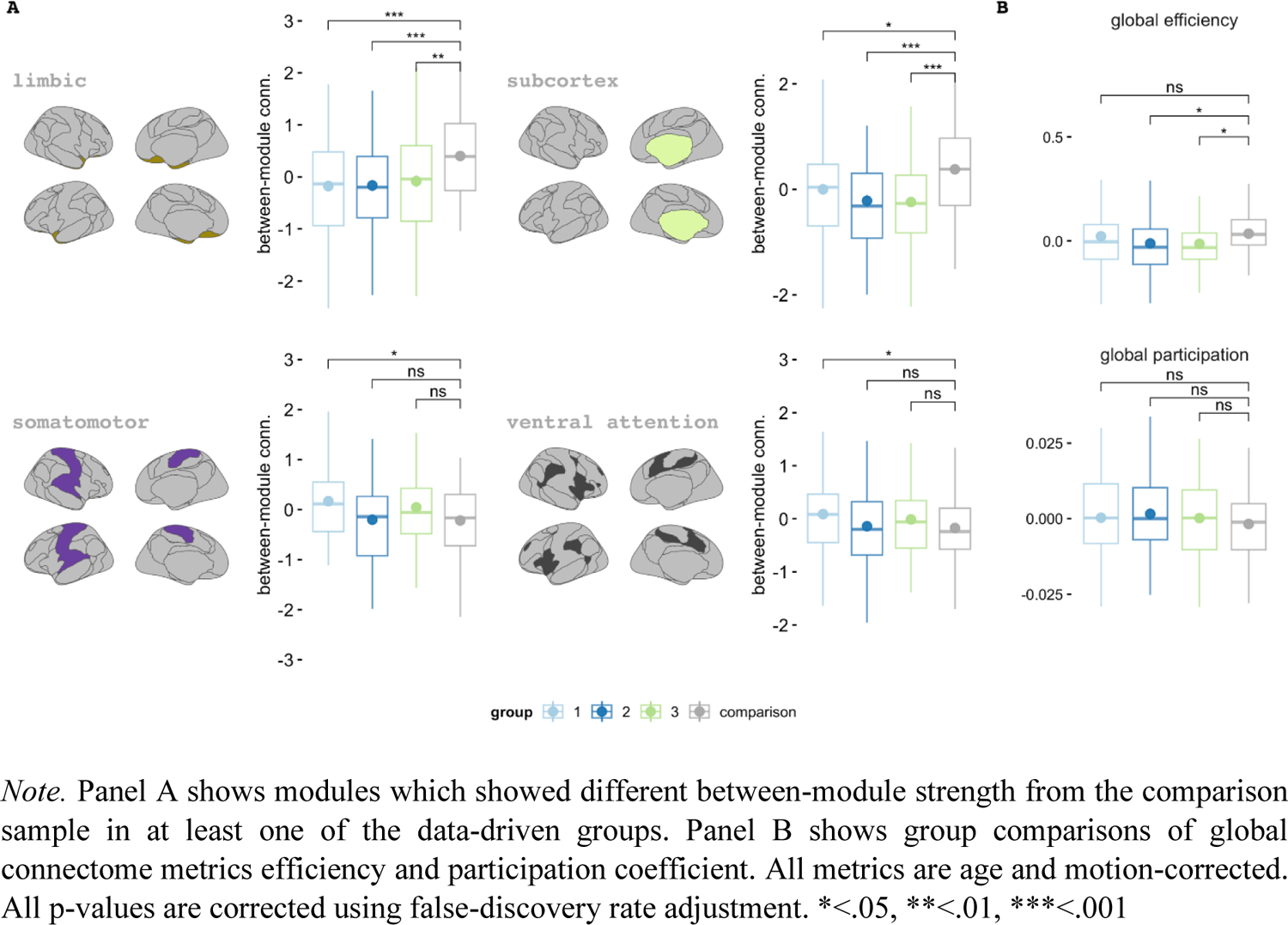
Group differences in modular and global organisation between each one of the three data-driven groups and the comparison group

Next, we identified 22 connector hubs that included regions within middle frontal gyrus, orbital gyrus, precentral gyrus, insula, cingulate gyrus, and the subcortex. The cross-validated balanced error rate across all models explored (k =1:5) suggested that the most accurate PLSDA model was the one with two components. The bootstrapped hub loadings onto the PLSDA components, which had 95% confidence intervals that did not cross zero, are shown in Figure 7. For the first component, robust loadings included regions mostly within the insula, as well as within the dorsolateral prefrontal cortex, superior temporal gyrus, and the basal ganglia. Permutation analyses suggested that relative to the non-referred comparison group, children with an inconsistent profile of predominantly task-based difficulties (Cluster 1) scored significantly lower on this component (p=.003). For the second component, all loadings that were shown to be robust in the bootstrapping procedure were frontal and parietal subregions. The second component distinguished participants with a profile of consistent difficulties across the executive function measures (Cluster 2) from the non-referred comparison group (p = .02), with component scores significantly higher in the comparison group.

**Figure 7.**
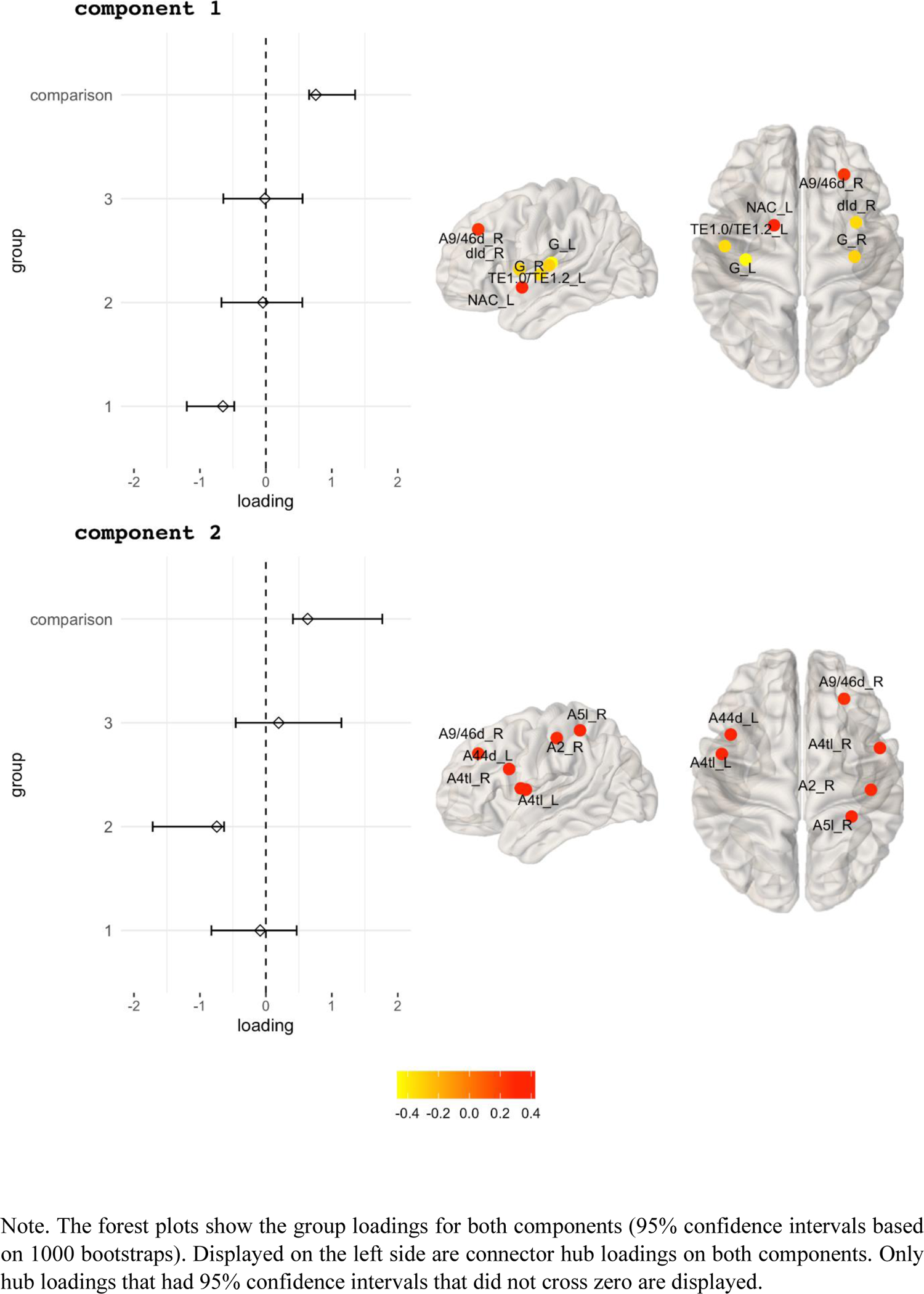
Bootstrapped component loadings of two components identified in the partial least squares discriminate analysis

## Discussion

This study used an unsupervised machine learning approach to map profiles of executive function among a large neurodivergent sample of children. Topographical maps were used to represent the profiles, which were subsequently carved into clusters representing homogeneous groups of children with similar profiles of executive function. Three clusters were identified, each with a distinct profile of executive function. The clusters could also be distinguished on measures of learning and mental health, but they cut across traditional neurodevelopmental diagnostic taxonomies: for the most part, they did not correspond to diagnostic categories or to dimensions of neurodiversity associated with common conditions such as ADHD or autism.

Three profiles of executive function were identified. One was characterised by difficulties that were consistent across the task- and rating-based measures of executive function (Cluster 2). The other two profiles were inconsistent across the two measurement types: children in Cluster 1 had close to age-expected levels on the rating-based measures but had difficulties on the performance-based tasks, while children in Cluster 3 had the opposite profile of close to age-expected performance on the task-based measures with pronounced difficulties on the rating-based assessments. In total, more than half the sample had an inconsistent profile. This provides further evidence that the two types of assessment (ratings and tasks) are dissociable, consistent with previous studies that have used latent variable or predictive validity approaches (e.g., Gerst et al., 2017; Nin et al., 2022; Soto et al., 2020; Tamm & Peugh, 2019; Toplak et al., 2013). The finding that a substantial proportion of our transdiagnostic neurodivergent sample showed little overlap between performance and rating based measures is also consistent with previous neurodevelopmental literature, which has shown that the two domains of executive function can be selectively affected within the boundaries of a single neurodevelopmental condition (e.g., those with ADHD, Biederman et al., 2008). Overall, the current findings support the idea that two types of measurement provide non-redundant information and should not be used interchangeably in the context of neurodevelopment.

### Cluster comparisons: behaviour

Children in Cluster 1, who were characterised by difficulties on the cognitive performance-based tasks with near to age-typical everyday executive function behaviours, were significantly younger than children in other two clusters, were less likely to have a diagnosed neurodevelopmental condition, and had fewer difficulties on dimensions of neurodiversity associated with ADHD, developmental language disorder and autism than children in the other two clusters. Girls were under-represented in this cluster. Children in this cluster performed more poorly on both learning measures than children in Cluster 3, who had the opposite profile of better task-than rating-based performance, and their maths scores were equivalent to those in Cluster 2 who had a consistent but poor executive function profile.

The broad profile of children in Cluster 1 suggests that diagnoses are less common among younger children, girls, and those with everyday behaviours that appear to be age-typical, despite poor performance on cognitive task-based measures of executive function and learning. This might reflect diagnostic systems that rely on observations of behaviour rather than performance on cognitive tests (the everyday behaviours of this group were almost age-typical), and in which children are not typically formally assessed for conditions such as ADHD until they are 7 years or older (American Psychiatric Association, 2013; Hoang et al., 2019; Root et al., 2019). Lower rates of diagnoses among this group, in which girls were over-represented, could also reflect differences in the outward characteristics of neurodivergent girls and boys: girls are more likely to use behavioural camouflaging strategies, appearing more able from others’ perspectives (Dean et al., 2017; Hiller et al., 2014; Hull et al., 2020), meaning they are less likely to receive a diagnosis (Dhuey & Lipscomb, 2010; Lockwood Estrin et al., 2021). Patterns of difficulties captured by parent/carer ratings on dimensions of neurodiversity associated with neurodevelopmental difficultes (e.g. ratings of inattention, restricted social interests etc.) reflected the same pattern as the diagnostic categories: fewer difficulties were present in children in this cluster relative to theose in the other clusters. This could again reflect masking / camouflaging, as well as socially constructed gender-biased or stereotypical views of boys as being disruptive (e.g., Sciutto et al., 2004). Together these findings underscore the need to include cognitive task-based assessments and female-representative characteristics in systems designed to identify children’s additional needs (see Guy et al., 2022 further discussion of this issue).

Children with the opposite profile of executive function, those in Cluster 3 with close to age-typical task-based performance and greater difficulties on the rating-based assessments, had significantly better performance on the learning measures than children in the other two clusters. This is consistent with a vast literature showing strong associations between performance on laboratory-based assessments of executive function and learning (Bull et al., 2008; Holmes et al., 2020; St Clair-Thompson & Gathercole, 2006). They also had significantly elevated self-reported internalising difficulties (anxiety and depression) compared to children in the other two clusters. This group also self-reported more internalising difficulties relative to the other two clusters, potentially suggesting a link between higher levels of executive function rating difficulties and mental health. Notably, this could not be entirely attributed to rater biases, given that mental health was self-rated and executive function ratings were provided by parents. The relationship between mental health difficulties and a profile of inconsistent and predominantly rating-based difficulties has previously been observed using different methods in this cohort (Williams et al., 2022). Others have suggested that rating-based difficulties occurring in the absence of any task-based performance problems arise through emotional rather than cognitive mechanisms (Vaidya et al., 2020; Williams et al., 2022), but longitudinal and/or causal evidence is needed to test this hypothesis.

Finally, those with the consistent profile of executive function difficulties had below-age-expected performance across both types of executive function assessments. Relative to the other clusters, they had similar ratings of executive function difficulties to children in Cluster 3, but significantly poorer performance on the task-based assessments than children in both other clusters. Their reading and mental health ratings were equivalent to those in Cluster 1, but their maths performance was significantly. Overall, they showed commonalities and differences with children in the other two clusters, which were not entirely attributable to measurement type (e.g., even though their rating-based executive function difficulties were equivalent to those in Cluster 3, they had significantly different mental health ratings).

### Cluster comparisons: neuroimaging

We observed both shared and cluster-specific patterns of neural white matter organisation differentiating data-driven groups from the comparison sample of neurotypical children who had not been referred for attention, learning, and/or memory difficulties. Children with either a consistent profile of executive difficulties (Cluster 2) and those with predominantly task-based difficulties (Cluster 3) had decreased global connectome efficiency relative to the comparison sample, but no group differences were observed in global participation. This was unexpected given previous reports of an association between modular segregation and executive function (Baum et al., 2017).

Looking at modular organisation, all three data-driven groups showed reduced connection strength within the limbic network and the subcortex relative to the non-referred neurotypical comparison group, with follow-up analyses showing that differences were driven by reduced inter-module connections. This is consistent with the idea that limbic and subcortical areas play a role in goal-directed cognition, and that differences in this circuitry are related to executive functioning in neurodevelopmental conditions (e.g., Arnsten & Rubia, 2012). The group with predominantly task-based difficulties (Cluster 1) also showed increased inter-module connection strength in the somatomotor and ventral attention modules, a pattern that was also observed in the PLSDA: the component of connector hubs differentiating this group from the non-referred neurotypical comparison group involved multiple insular regions of the somatomotor and ventral attention modules. Somatomotor and ventral attention networks undergo substantial restructuring over the course of development, becoming more structurally segregated with age (Baum et al., 2017; Grayson & Fair, 2017). Observing differences in this network among children with predominantly task-based difficulties is consistent with their implicated role in task-based executive functions (Reineberg et al., 2015). Additionally, several of the hubs which loaded robustly on the PLSDA component that differentiated this group from the neurotypical sample shared perception as a behavioural metadata label within the BrainMap Database (www.brainmap.org/taxonomy).

Children with a profile of consistent difficulties across the task- and rating-based measures (Cluster 2) were uniquely distinguished from the non-referred comparison group through a component of frontal and parietal connector hubs identified through the PLSDA. This is consistent with decades of research that has established a role for frontal and parietal regions in supporting executive function (see Friedman & Robbins, 2022). Exploratory cross-checking of the behavioural metadata labels for the hubs robustly loading on this component within the BrainMap Database suggested that many of them were functionally implicated in action execution.

Finally, considering other similarities, the two components identified by the PLSDA that differentiated the two groups with task-based executive function difficulties (Cluster 1 and 2) from the non-referred comparison sample had a single shared robust loading. This region, located in the right dorsal area 9/46 within the middle frontal gyrus, is functionally associated with task-based executive function performance on measures of working memory and attention (e.g., Jung et al., 2022).

### Limitations

The measurement types used were limited by those available in the cohort data. As such, it is possible, for example that stronger links between the everyday manifestations of executive function difficulties captured by the rating scales and learning could have been detected if we did not have to rely on measures of learning administered under optimal testing conditions, but instead had access to school-based measures of academic achievement that reflect learning and assessment in everyday conditions. Methodologically, the combination of a continuous multidimensional mapping method with a data-driven clustering algorithm means that inevitably some children will sit close to the cluster boundaries within the map. In terms of the neuroimaging results, it is important to acknowledge that the use of multimodal neuroimaging and larger samples may reveal further and equally important differences across groups, which we consider an important future direction. Finally, the exploratory nature of this investigation should be noted. We encourage further efforts to replicate and extend these findings to better understand the diversity of executive function profiles among neurodivergent individuals.

## Conclusion

In summary, we used a machine learning approach to map the executive function profiles of large neurodevelopmentally neurodivergent sample. We identified three distinct profiles that were validated by differences across measures of learning, mental health, and neural white matter organisation. These data add to the growing evidence base for two conceptualisations of executive function: one that reflects higher-order cognitive skills and can be measured by cognitive tasks (Miyake et al., 2000), and the other that reflects the application of cognitive control in everyday contexts and can be measured by rating scales (Doebel, 2020). The majority of our neurodivergent sample had inconsistent profiles across the two domains, suggesting that researchers and practitioners should not use these two types of assessments interchangeably in the context of neurodevelopment. They should instead be used together in a complementary manner.

## Data Availability

Data available from https://calm.mrc-cbu.cam.ac.uk/

## Acknowledgements

The CALM Team includes lead investigators Duncan Astle, Kate Baker, Susan Gathercole, Joni Holmes, Rogier Kievit and Tom Manly. Data collection is assisted by a team of researchers and PhD students that includes Danyal Akarca, Joe Bathelt, Madalena Bettencourt, Marc Bennett, Giacomo Bignardi, Sarah Bishop, Erica Bottacin, Lara Bridge, Diandra Brkic, Annie Bryant, Sally Butterfield, Elizabeth Byrne, Gemma Crickmore, Edwin Dalmaijer, Fanchea Daly, Tina Emery, Laura Forde, Grace Franckel, Delia Furhmann, Andrew Gadie, Sara Gharooni, Jacalyn Guy, Erin Hawkins, Agnieszka Jaroslawska, Sara Joeghan, Amy Johnson, Jonathan Jones, Rebeca Ianov-Vitanov, Christian Iordanov, Silvana Mareva, Jessica Martin, Alicja Monaghan, Elise Ng-Cordell, Sinead O’Brien, Cliodhna O’Leary, Joseph Rennie, Andrea Santangelo, Ivan Simpson-Kent, Roma Siugzdaite, Tess Smith, Stephani Uh, Maria Vedechkina, Francesca Woolgar, Natalia Zdorovtsova, Mengya Zhang. The authors wish to thank the many professionals working in children’s services in the South-East and East of England for their support, and to the children and their families for giving up their time to visit the clinic. The CALM dataset is not yet available as the study is still ongoing. The data will be made available via managed open access once the study is complete. S.M. and J.H. conceived and designed the study, analysed the data, and wrote and edited the manuscript. The CALM team collected the data. Dr Danyal Akaca pre-processed the neuroimaging data. All authors approved the final version. Ethical approval was granted by the National Health Service. Parents/caregivers provided written consent and child verbal assent was obtained. The authors have declared that they have no competing or potential conflicts of interest.

## Supplementary materials

**Figure S1.**
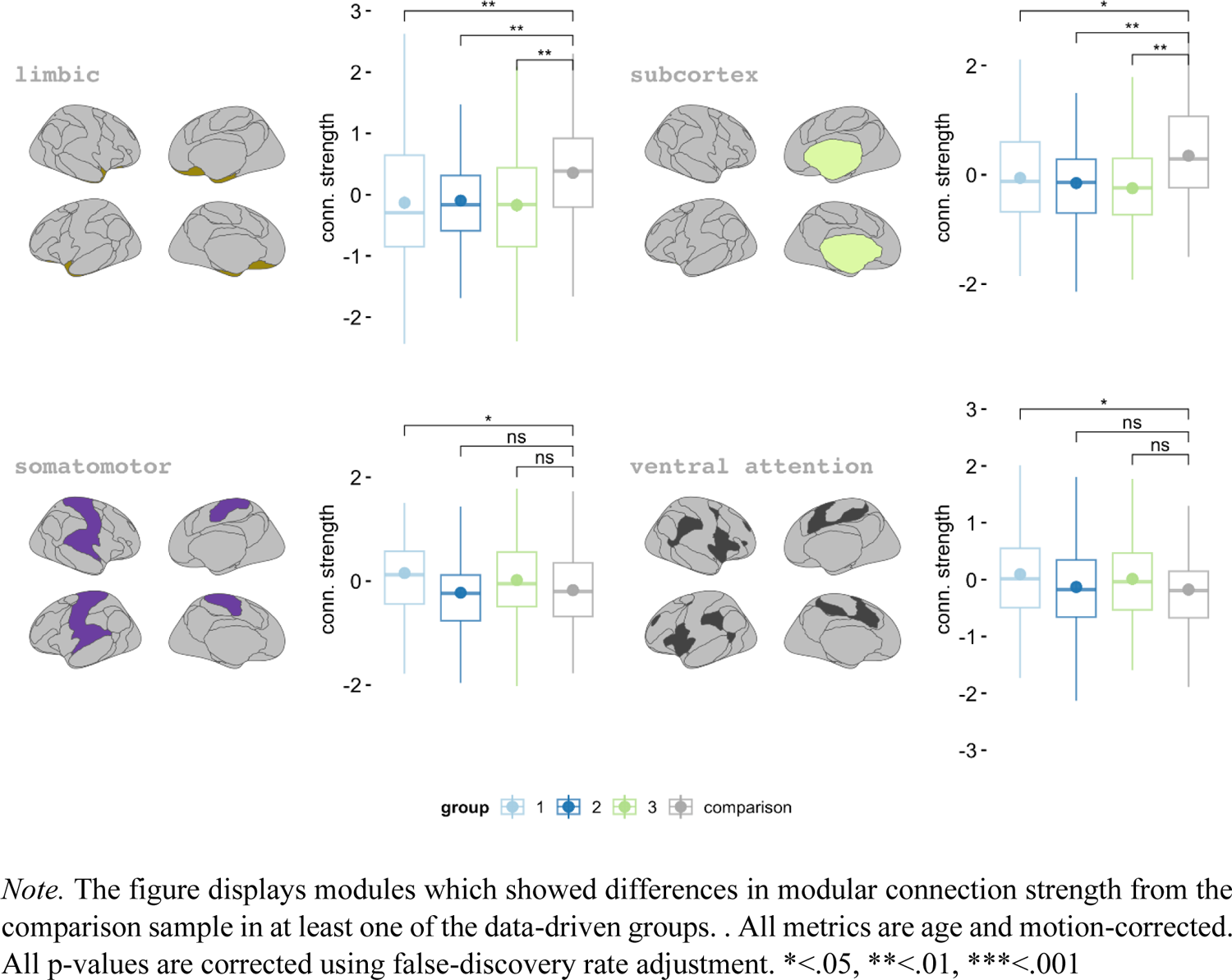
Group differences in modular strength between each one of the three data-driven groups and the comparison group

